# When does contacting more people lessen the transmission of infectious diseases?

**DOI:** 10.1101/2021.08.09.21261796

**Authors:** Bernardo A. Mello

## Abstract

A primary concern in epidemics is to minimize the probability of contagion, often resorting to reducing the number of contacted people. However, the success of that strategy depends on the shape of the dose-response curve, which relates the response of the exposed person to the pathogen dose received from surrounding infected people. If the reduction is achieved by spending more time with each contacted person, the pathogen charge received from each infected individual will be larger. The extended time spent close to each person may worsen the expected response if the dose-response curve is concave for small doses. This is the case when the expected response is negligible below a certain dose threshold and rises sharply above it. This paper proposes a mathematical model to calculate the expected response and uses it to identify the conditions when it would be advisable to reduce the contact time with each individual even at the cost of increasing the number of contacted people.

## Introduction

Within the recent effort on understanding the evolution of Covid-2019, an essay by S. Mukherjee [1] poses two relevant questions regarding the initial viral dose that a susceptible individual receives from an infected person:

*Question 1:* Does the initial dose affect the probability of infection?

*Question 2:* Does the initial dose affect the severity of the disease?

Though not asked by him, a pertinent question when investigating the propagation of disease is

*Question 3:* Does the initial dose affect the subsequent infectiousness of the exposed person?

These questions are related to two usual goals of epidemics management: reducing the spread of diseases and the severity of the symptoms.

A key concept when answering the questions is the dose-response curve *π*(*q*), which estimates the expected severity of the outcome as a function of the pathogen dose *q*. This curve describes the probability or the expected severity of an outcome, such as infectiousness, immunity, contagion, mild symptoms, severe symptoms, and death. When investigating the spread of the disease, infectiousness and immunity are probably crucial information, but it is also relevant to evaluate the symptoms and the death probably. The dose-response curve was recently employed to describe how the protection against COVID-19 from wearing masks depends on the environmental virus concentration [2].

It is difficult to answer the questions because it is often impossible to measure the initial dose directly. There are relatively few papers focusing on these questions and even fewer trying to find the dose-response curve, *π*(*q*). Notwithstanding the difficulties, question 1 has been explored for hematopoietic necrosis virus in trouts [3], antrax [4,5] cytomegalovirus [6,7], herpes simplex virus-2 [8], HIV-1 [9–11], and SARS-Cov-2 [12]. Question 2 was addressed for SARS-Cov-2 in [13]. Both questions were indirectly addressed by exploring the microscopic dynamics of infection by poliomyelitis viruses [14], Moloney sarcoma virus [15,16], and herpes simplex virus-2 [17].

It is common for a person not to be able to avoid sharing limited space with other people. Some examples are hospitals, transportation, classrooms, restaurants, and workplaces. Still, in certain cases, the number of distinct people approached by each person can be reduced or increased. For instance, students can be directed to change or keep places at each new class [18]. Staff could alternate the patients and clients attended in hospitals and restaurants. Rules could be applied to pedestrian traffic [19]. Forced ventilation could be used in a closed environment to homogenize the pathogen concentration, playing a role similar to altering the distance between people [20].

If a person encounters many different people but stays for a short time with each of them, he or she will be subject to a low exposition when meeting an infected person. Conversely, if he or she encounters fewer people but stays longer with each person, the chance of encountering a contagious person is lower; but the contamination received from each infected person is higher. As it will be seen, even if the mean exposition is the same in both cases, the standard deviation is different, and this difference can play an essential role in the expected response.

This paper presents a simple mathematical model to quantify the expected outcome of changing the number of contacts. It depends on four quantities:

*γ* Fraction of infectious people in the population.

*N*_*c*_ Number of contacted people, understood as the number of people that got close enough to transmit the pathogen.

*κτ/Q* Ratio between the utmost pathogen charge (*κτ*), which would be received if every person met was infected, and the charge expected to generate 50 % of the maximum response (*Q*).

*h* Parameter that controls the concavity of the dose-response curve for low doses, with the form

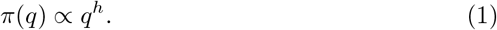

for small values of *q*.

In the *Materials and Methods* section, we formulate the model, demonstrate the importance of *π*(*q*)’s concavity with a normal distribution of pathogen dose, and apply it to a population of infected people, best described by the binomial distribution of pathogen doses. The numerical evaluation of the model is presented in the *Results* section and analyzed in the *Discussion* section.

## Materials and methods

### The response curve

We will consider a person who stays close to other *N*_*c*_ people while engaged in certain activity for a period of time *T*. The equivalent contact time of that person is defined as

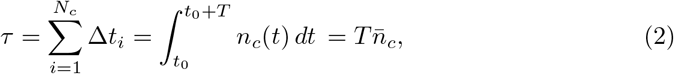

where Δ*t*_*i*_ is the time spent close to person *i* and *n*_*c*_(*t*) is the number of nearby people at time *t*. The equivalent contact time is equal to the total time, *T*, multiplied by the temporal average of the number of nearby people, 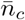.

The binary variable *γ*_*i*_ defines the infectious state of the person *i*, with the value 0 for non-infectious and 1 for infectious. The fraction of infectious people in the population of size *N*_pop_ ≥ *N*_*c*_ is 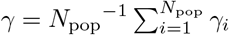. As a simplifying hypothesis, we assume that nearby infectious people transmit the pathogen to the exposed person with the constant rate *κ* and that transmission is not possible from afar. Therefore, the charge received from the person *i* is

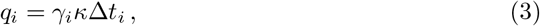

and the total charge received is

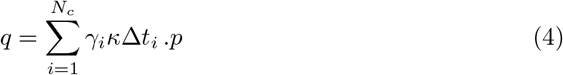

Brouwer at al [21] demonstrated that the concavity of the response curve for low doses plays a crucial role in the transmission models of environmentally mediated infectious diseases. Among the curves explored by the authors, only the Hill and the Weibull distributions allow changing the concavity. As discussed in S1 Appendix, these are distinct curves, but their parameters can be adjusted to achieve partial superposition of one over the other within a curve’s sector. As it will be shown, most of the intriguing results in this paper depend on the behavior of the curve with small values of *q*. S1 Appendix provides information that allows estimating the values of the parameters of the Weibull distribution that shall produce results similar to the Hill curve in certain limits.

This work uses the Hill curve, but it is reasonable to assume that similar behaviors would result with any function *π*(*q*) that possesses the following four properties: (*a*) it is zero for *q* = 0; (*b*) it increases monotonically with *q*; (*c*) it approaches a value less than or equal to one as *q*; → ∞; (*d*) its concavity near zero can be adjusted as the parameter *h* in Eq. (1).

We write the dose-response curve as

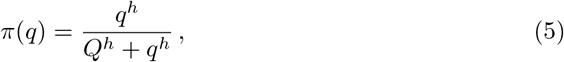

where the half response charge, *Q*, is the charge at which the expected response is half of the maximum probability, reached when *q* → ∞. When *h >* 1, this curve has an inflection point, defined by 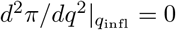, at

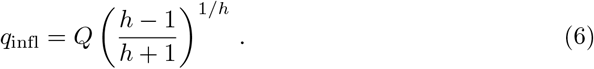

As shown in Fig. 1, when *h* ≤ 1, the curve is concave everywhere, and when *h >* 1, the curve is convex at the left side of the inflection point and concave at the right side. For *h* = 1, the expected response is proportional to the pathogen charge when this charge is low. For *h <* 1, minute charges have a high expected response. For *h >* 1, the expected response is negligible below a pathogen charge threshold.

**Fig 1.**
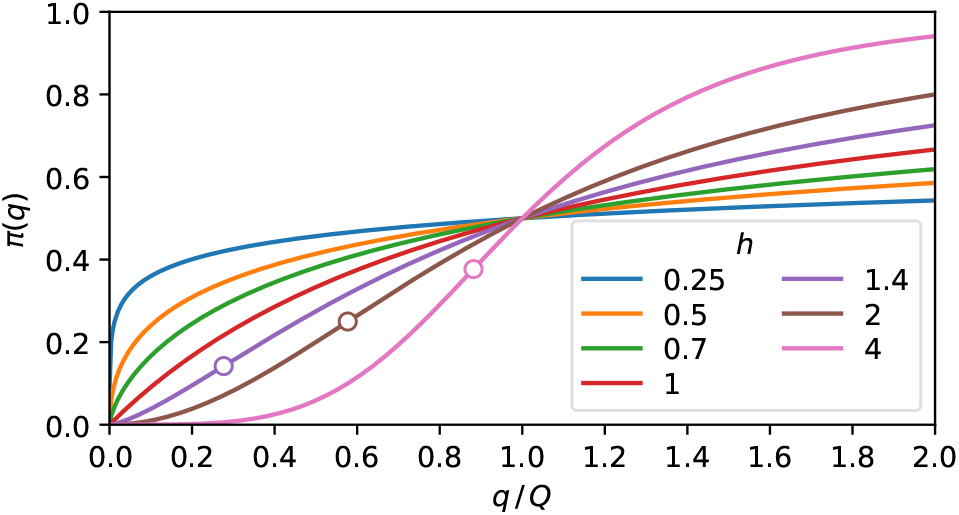
The expected response. Plot of Eq. (5) for some values of *h*. The curves are concave for *h* ≤ 1. When *h >* 1, the convex and the concave parts are, respectively, at the left and right sides of the inflection point, marked as a circle.

### The concavity of the response curve

When a group of people is submitted to the conditions described in the previous section, with the probability *P* (*q*) of receiving the charge *q*, its mean charge and variance are

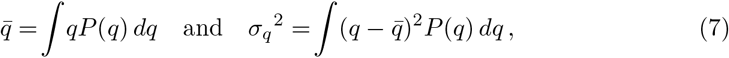

and the expected response of this population is

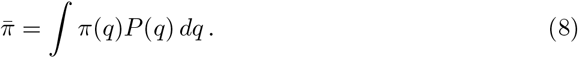

Since the response curve is not a linear function and the population covers a range of pathogen charges, the population’s expected response is not equal to the expected response of the mean population charge, i.e., 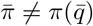.

If the charge probability distribution is strongly peaked around 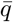, with 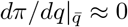, *π*(*q*) may be approximated as a Taylor expansion up to the second-order around 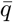 in Eq. (8), resulting in

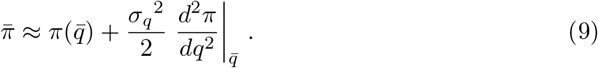

This expression indicates that if two symmetric distributions of pathogen doses have the same mean value, the wider one will have a higher expected response if the second derivative is positive. Thus, broadly speaking, a wider population will have a higher expected response if *π*(*q*) is convex in the vicinity of 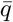, and a lower expected response for concave vicinity.

Figure 2 highlights the dependence of the population’s expected response on the concavity of the response curve and the population exposure distribution. If the population exposure is strongly peaked, the population’s expected response is very close to the value of the response curve at the mean population charge, as shown in Fig. 2A. On the other hand, if the population charge is too diverse, the population’s expected response falls unmistakably above or below the response curve, depending on the concavity, as can be seen in Fig. 2B. According to Eq. (9), the expected response should sit on the curve for the middle distributions of Fig. 2, since *d*^2^π / *dq*^2^ = 0 at their centers. The difference observed in the percentages of Fig. 2B manifests the inadequacy of Eq. (9) for large values of *σ*_*q*_.

**Fig 2.**
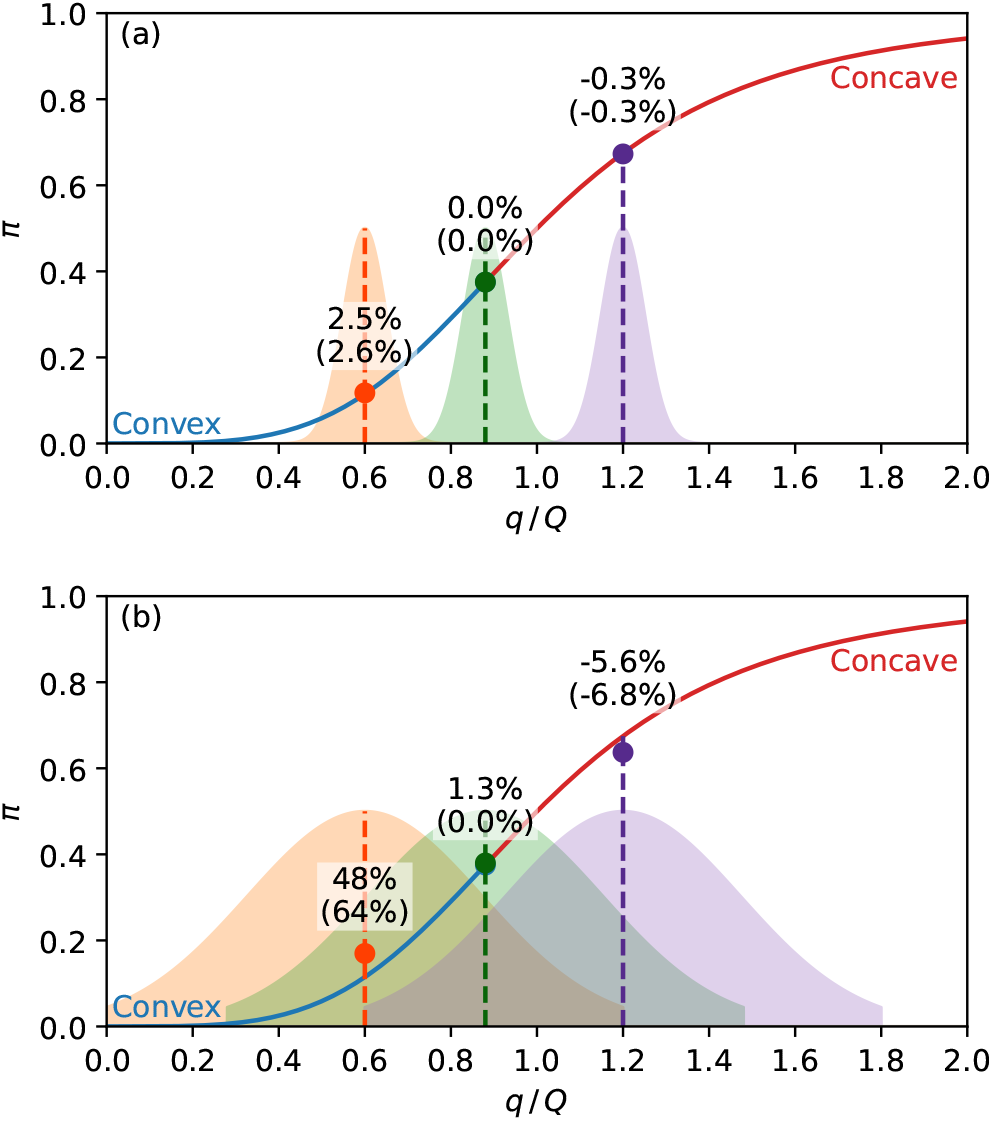
Expected response of a population. The solid line is Eq. (5) with *h* = 4, with the convex part in blue and the concave part in red. The shaded areas are the population distributions of the pathogen charge in arbitrary units, with the middle distribution centered at the inflection point. The circles are the average expected response of each distribution, given by Eq. (8). The percentages are the relative difference of the average expected response from the response function at the center of the distribution 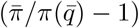. The percentages between parenthesis are the same quantities calculated with the approximation Eq. (9). A: Narrow charge distributions (*σ*_*q*_ = 0.05) result in the population’s expected response close to the value of the response curve at the mean charge. B: For broader distributions (*σ*_*q*_ = 0.25), the population’s expected response is above or below the response curve, respectively, at the convex and concave parts of the curve.

### Uniformly divided contact time

The total exposure, Eq. (4), is a sum of *N*_*c*_ equally distributed random quantities *γ*_*i*_*κ*Δ*t*_*i*_. We will now consider the situation where *N*_*c*_ and Δ*t*_*i*_ are, respectively, multiplied and divided by the same factor. This operation preserves the value of 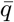 and, by the Central Limit Theorem, makes standard deviation proportional to 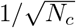.

A simple concrete situation that exhibits that behavior is an individual with an equivalent contact time *τ*, which is equally divided among *N*_*c*_ other people, resulting in the same time Δ*t*_*i*_ = *τ/N*_*c*_ spent near each of them. From Eq. (4), the pathogen charge received by this person is

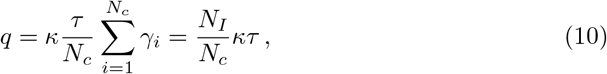

where *N*_*I*_ is the number of infected people met. The utmost pathogen charge, *κτ*, is the charge a person would receive if all people met were infected, i.e., if *N*_*I*_ were equal to *N*_*c*_. Instead, the number of infected people follows the binomial distribution,

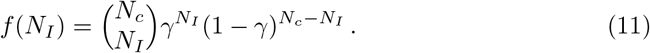

From the mean value, 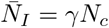, and the standard deviation, 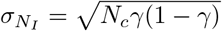, of the binomial distribution, the mean value and the standard deviation of the total charge received may be obtained,

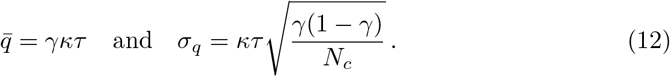

The standard deviation presents the 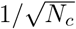 behavior mentioned above.

In the following lines, we obtain asymptotic expressions of 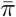 at the limits *N*_*c*_ *→* 0 and *N*_*c*_ → ∞. If the mean number of encounters with infected people is low enough, *N*_*I*_ = *γN*_*c*_ ≪ 1, most contacts with infected people will be with just one person. From Eq. (10), the charge of such encounter is *κτ/N*_*c*_, and the following approximation is valid for the expected response

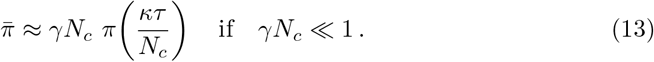

This expression is exact for *N*_*C*_ = 1. By handing *N*_*C*_ as a real number, the maximum of Eq. (13) is approximately defined by 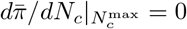, resulting in

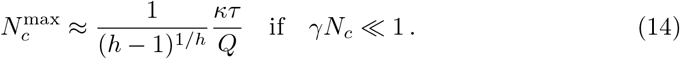

At the limit *γN*_*c*_ ≫ 1, the binomial distribution is strongly peaked around the mean value, *γκτ*, and the corresponding expected response is

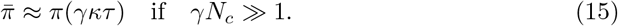

The value of the Eq. (14) is not real for *h <* 1, and 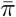 given by Eq. (8) grows monotonically from *N*_*c*_ = 0 to *N*_*c*_ = ∞ For *h >* 1, the existence of a maximum 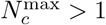 requires 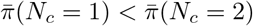, and from Eq. (13) results in

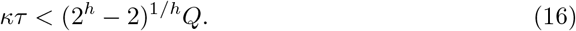

If this condition is satisfied, there is a maximum at 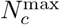 and two minima, at *N*_*c*_ = 1 and *N*_*c*_ = ∞. By substituting *N*_*c*_ = 1 in Eq. (13) and making it equal to Eq. (15) with *N*_*c*_ → ∞ we obtain

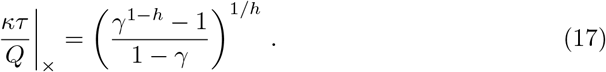

At the left and the right of the this quantity, the global minimum is, respectively, at *N*_*c*_ = ∞ and *N*_*c*_ = 1.

## Results

The analytical approximations, Eq. (9), Eq. (13), and Eq. (15), help understand the qualitative properties of the mean response. This section presents the exact numerical calculation of 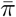 from Eq. (8), with the Gaussian distribution and the binomial distribution, Eq. (11).

The Gaussian population’s expected response illustrated by Fig. 2 suggests that the contagion rate may be reduced by changing the distribution at the right of Fig. 2A to 2B, i. e., it is possible to reduce the expected response by making the distribution wider at the concave part of the response curve. The the standard deviation increase leading to a reduction in 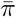 illustrates this behavior in Fig. 3A, Fig. 3B, and for 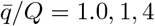 and 2.0 of Fig. 3C. The response of the distributions at the left of Fig. 2, which is lower in Fig. 2A than in 2B, demonstrates the reverse behavior. I. e., making the distribution thinner reduces the response at the convex part of the function, as illustrated by the curves with 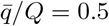 and 0.7 of Fig. 3C.

**Fig 3.**
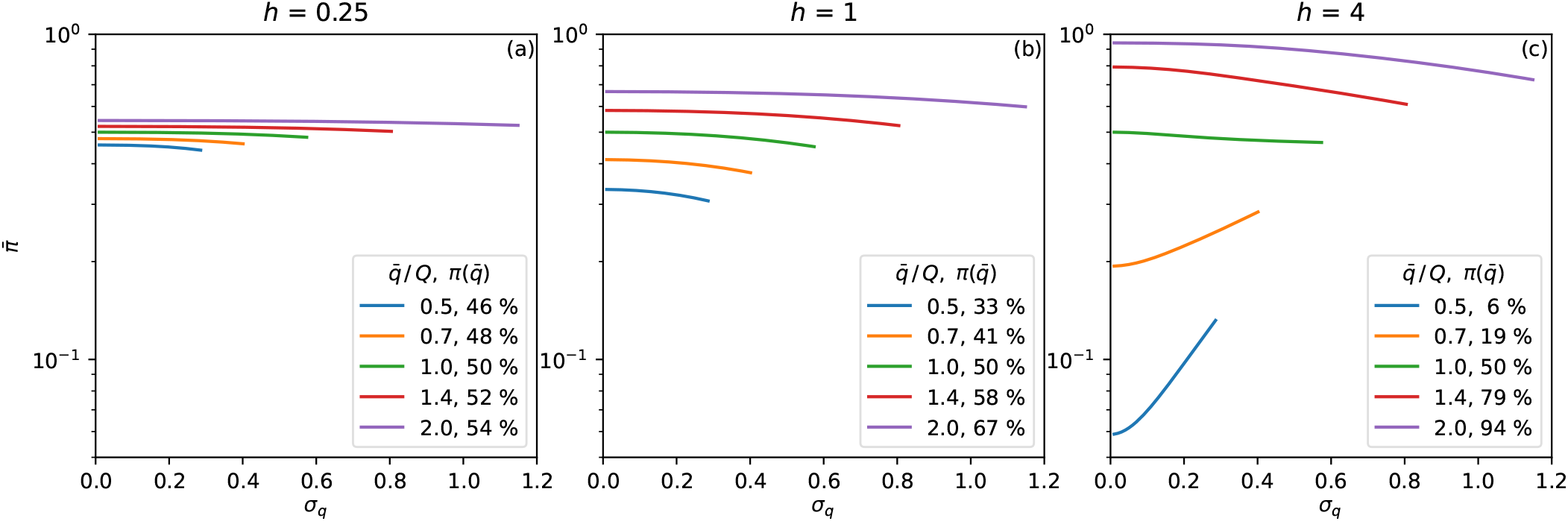
Gaussian distribution. Expected response as a function of *σ*_*q*_, Eq. (7)-(8). The symmetry of the distributions is preserved by truncating them at 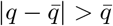. The curves are plotted up to the highest *σ*_*q*_ allowed by the truncation for each value of 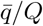. The value of the curve with *σ*_*q*_ = 0 in Eq. (9) is shown as the percentage in the legend. A: *h* = 0.25, B: *h* = 1, C: *h* = 4.

Dividing the contact time with more people but spending proportionally less time with each of them results in a thinner distribution with the same mean value. The behavior demonstrated by the distributions at the left of Fig. 2 indicates that this would be advisable for convex dose-response curves.

While the Gaussian distribution is a standard choice, more realistic pictures require describing how the person divides the contact time among several people. The minimalist model discussed above results in the binomial distribution, Eq. (11), investigated below.

We will determine the number of contacted people, *N*_*c*_, that minimizes the binomial distribution’s expected response. That distribution emerges if a person can choose how many people to meet, spending with each person a time inversely proportional to the number of people met. For *h* ≤ 1, 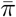 is a monotonically increasing function of *N*_*c*_, as can be seen in S6 Fig. Therefore, for *h* ≤ 1, as few people as possible should be contacted to reduce the expected response.

In Fig. 4 we can see 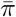 for the binomial distribution as a function of *N*_*c*_ for several combinations of *h, γ*, and *κτ*, with *h >* 1. The values of 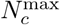 predicted by Eq. (14), represented by black dashed lines in Fig. 4, are in good agreement with the exact calculation assigned as hollow circles. The value of 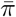 with *N*_*c*_ → ∞ is plotted as the rightmost point of each function.

**Fig 4.**
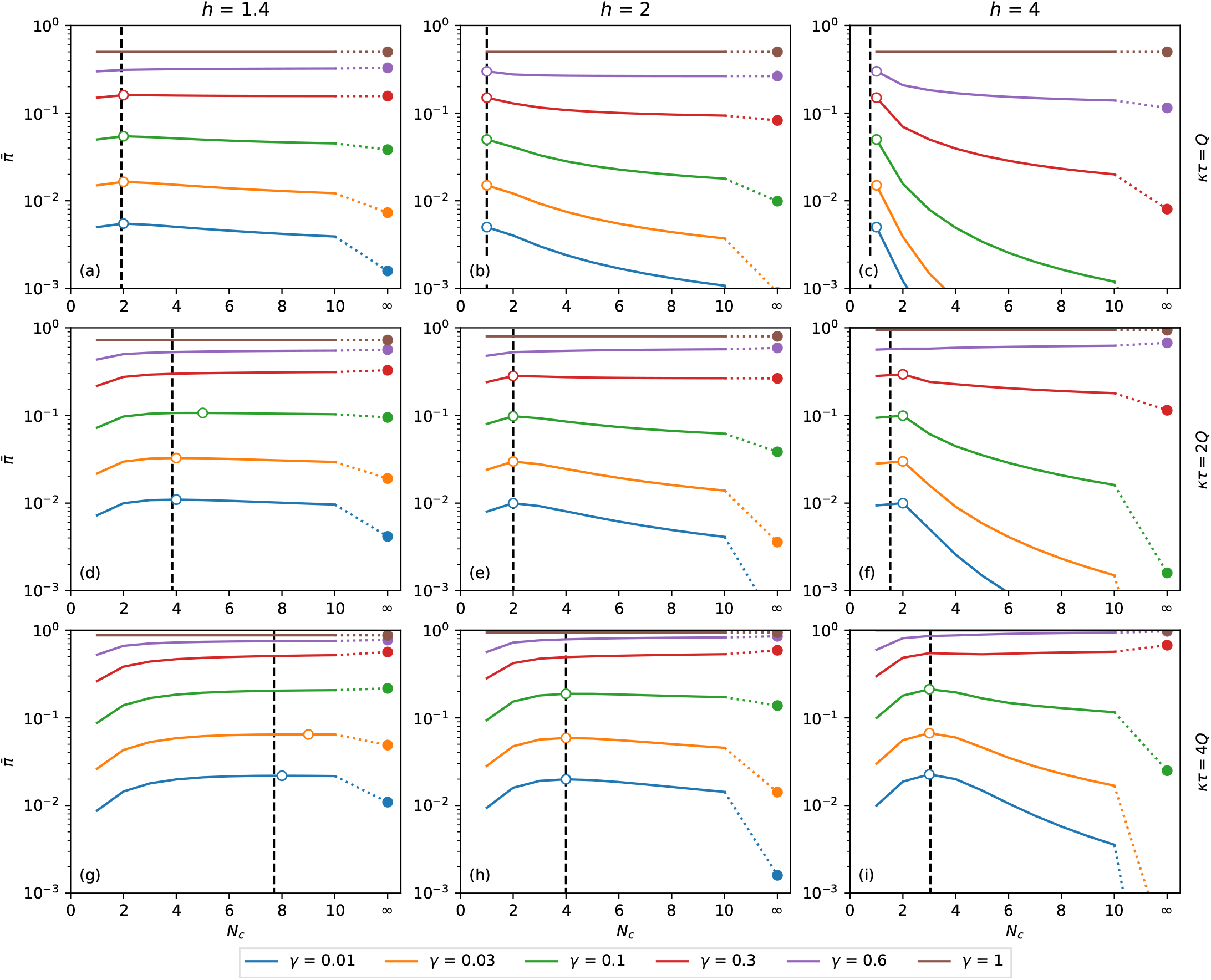
Binomial distribution. Expected response as a function of *N*_*c*_. It is calculated as a function of the number of contacts for the binomial distribution of pathogen charge, Eq. (10)-(11) applied to Eq. (8). Although *N*_*c*_ is an integer variable, the functions are shown as lines to make the plots less bulky. The dashed vertical lines are the points of maximum predicted by the approximation Eq. (14). The hollow circles are the points of maximum of each combination of *h, κτ*, and *γ*. The filled circles are the values of 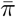 at *N*_*c*_ → ∞, calculated as *π*(*γκτ*), Eq. (15). A: *κτ* = *Q, h* = 1.4, B: *κτ* = *Q, h* = 2, C: *κτ* = *Q, h* = 4, D: *κτ* = 2*Q, h* = 1.4, E: *κτ* = 2*Q, h* = 2, F: *κτ* = 2*Q, h* = 4, G: *κτ* = 4*Q, h* = 1.4, H: *κτ* = 4*Q, h* = 2, I: *κτ* = 4*Q, h* = 4.

In order to minimize the expected response, it may be necessary to increase or reduce the number of contacts, depending on the values of *h, γ*, and *κτ/Q*. From Eq. (16), *N*_*c*_ = 1 is a maximum of 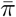 if *κτ* ≤ 0.78*Q*, 1.5*Q*, and 1.97*Q*, respectively, for *h* equal to 1.4, 2, and 4. As shown in Fig. (4), if this condition is satisfied, then 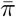 is monotonically decreasing with *N*_*c*_, and the minimum expected response is obtained when the number of contacted people is maximized.

The picture is more complex if with finite 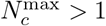. Figure 5 summarizes the information required to determine how to reduce the expected response in each case. The number of contacts that maximizes the expected response, 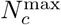, is plotted as a function of *κτ/Q* in Fig. 5A-C for some combinations of *h* and *γ*. For low exposure time (*κτ/Q →* 0), the maximum is at 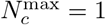 and grows with *κτ/Q* in steps of unitary height. When *κτ/Q* reaches a specific value, 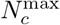 becomes infinite, signaling that, for *κτ/Q* larger than that value, 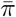 is a monotonically growing function of *N*_*c*_, and that the number of contacts must be minimized to reduce the expected response.

**Fig 5.**
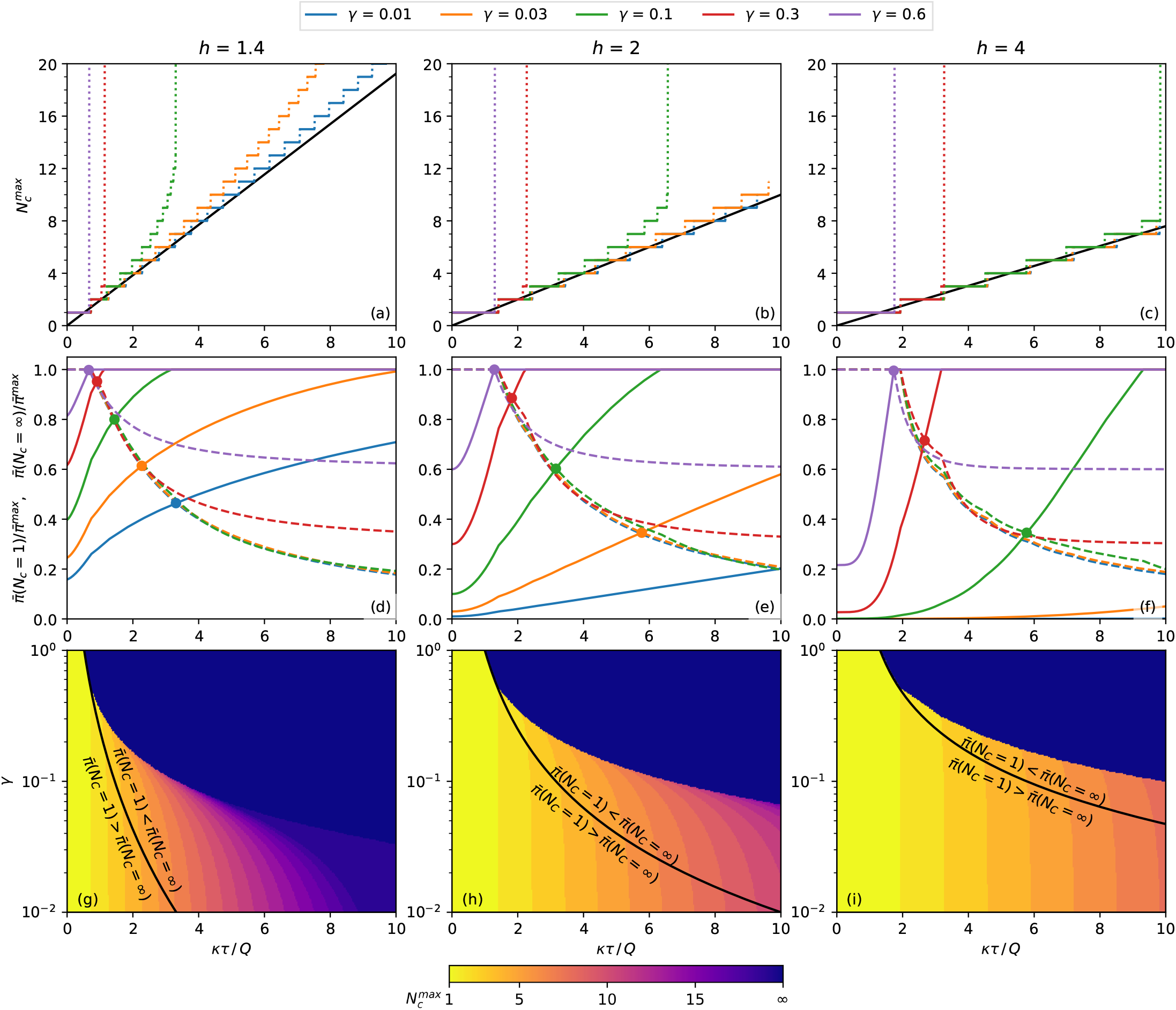
Properties of the population’s expected response. (a)-(c) Number of contacts that result in a maximum probability of contagion for plots such as Fig. 4. The continuous black line is the value predicted by Eq. (14). (a) *h* = 1.4, (b) *h* = 2, (c) *h* = 4. (d)-(f) The dashed lines and the continuous line are the expected response, respectively, at *N*_*c*_ = 1 and *N*_*c*_ = ∞, both relative to the maximum expected response, 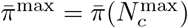. The circles mark the crossings of 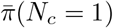 and 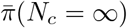 with the same *γ*. (d) *h* = 1.4, (e) *h* = 2, (f) *h* = 4. (g)-(i) Equation (17), plotted as black lines, separates the regions of the *γ*-*κτ/Q* phase space where the inequalities written on each side of the curve are observed. The color represents the 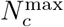 of each combination of *γ* and *κτ/Q*. (g) *h* = 1.4, (h) *h* = 2, (i) *h* = 4.

If 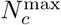 is finite, it is necessary to inspect the boundary values of the expected response, 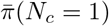 and 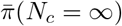. These quantities are plotted in Fig. 5D-F, and their crossing point, described by Eq. (17), are marked by circles. At the left of the crossing point, 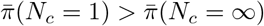 and at the right of the crossing point, 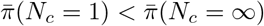. To reduce the expected response of each combination of *h* and *γ*, the number of contacts should be infinite for *κτ/Q* less than crossing value, and the number of contacts should be 1 for *κτ/Q* bigger than the crossing value. The black lines of Fig. 5G-I represent these frontiers as a continuous function of *γ*.

The above conclusion is only valid if *N*_*c*_ can be freely chosen in the interval 1 ≤ *N*_*c*_ ≤ ∞ (or that the maximum value of *N*_*c*_ is big enough to be considered infinite). With other lower or upper limits for *N*_*c*_, a specific calculation may be needed. However, these calculations are unnecessary if the minimum allowed number of contacts is greater than 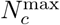, when the number of contacts should always be maximized. Similarly, if the maximum allowed number of contacts is less than 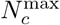, the number of contacts should always be minimized. The color plot in Fig. 5G-I represents the value of *N*_*c*_ as a function of *γ* and *κτ/Q*.

## Discussion

This paper describes how people sharing a limited space in an epidemic respond to the pathogen exposition from their companions. They cannot avoid being close to each other but can change the time spent near each person. The mathematical model supposes five simplifying hypotheses: (*a*) the number of contacted people changes but the number of nearby people averaged on time is constant, (*b*) the same time is spent near every person approached, (*c*) each contacted person can be infectious or not, with no intermediate states, (*d*) the pathogen is received from nearby infectious people at constant and identical rates, (*e*) distant people do not transmit the pathogen.

These hypotheses lead to a simple solution and make it evident what are the main parameters controlling the results. However, more information is necessary to build a detailed model, for example, regarding the viral shedding dynamics [22,23].

Under the above hypothesis, reducing the number of people met leads to an increase in the standard deviation, as expressed by the dependence on 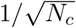 of Eq. (12). Most curves of 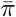 in Fig. 3 decrease monotonically with *σ*_*q*_, implying that cutting down the number of contacted people, which increases *σ*_*q*_, diminishes the response. However, when *h >* 1, small values of 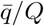 generate monotonically growing functions (Fig. 3C with 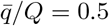 and 0.7), originating a counter-intuitive behavior: decreasing the number of contacted people increases the standard deviation and the expected response. The narrower distribution of the received pathogen charge is the cause of this startling *evenness curtailing* in the expected response, observed in the convex part of the response curve, Fig. 2.

The following argument clarifies the origin of the evenness curtailing. When the distribution of received doses is wide, a large fraction of the population is exposed to doses much higher or much lower than the average. If the response curve is convex, the increase in the response of the overexposed people will be more significant than the decrease in the response of the underexposed people, leading to a positive net effect. The wider the distribution, the stronger the average response.

As expressed by Eq. (9), the responsible for evenness curtailing is not the reduction in the average change but the change in the standard deviation. Increasing the number of contacts while keeping the total contact time constant homogenizes the exposition over a larger set of individuals, reducing the standard deviation. Therefore, even if the simplifying hypotheses (*b*)-(*e*) are not assumed, increasing contacts should still lessen the response in certain situations.

Unfortunately, little is known about a central aspect of this phenomenon: the parameter *h*. In the S2 Appendix, we review some experimental works that provide information suitable for estimating the value of *h*. By fitting Eq. (5) to the experimental data, we find *h* ranging from 1.12 to 2.29.

As a concrete example of applying the approach presented here, let us imagine a response curve with *h* = 2 and *γ* = 3 % of the population transmitting the pathogen. This percentage is less than the fraction of exposed or infected individuals since these patients may not be shedding the pathogen. From Fig. 5E, if *κτ* ≲ 5.7*Q*, the mean response will be lower for *N*_*c*_ → ∞ than for *N*_*c*_ = 1. Suppose that a worker must spend 8 hours in a workplace, sharing a workstation with three colleagues, resulting in *τ* = 24 h of equivalent contact time. Let us also assume that a 12 h exposition to an infectious person, adding up to the charge *Q* = 12*κ*, leads to a contagious probability of 50 %. Therefore, the worker’s utmost pathogen charge is *κτ* = 2*Q*. If the workers keep their place for the whole shift, they will contact the same three individuals through their shift. In this case, *N*_*c*_ = 3 and from the data used to plot Fig. 4E we obtain a contagion probability of 28 %. If the workers change places every 4 hours, *N*_*c*_ = 6 and the contagion probability drops to 19 %. By changing place every 2 hours, *N*_*c*_ = 12 and the contagious probability is 12 %. By comparing Fig. 4B and Fig. 4E, we conclude that the reduction achieved by increasing *N*_*c*_ would be more robust if the fraction of infectious people, *γ*, or if the utmost pathogen charge, *κτ* were lower. Increasing the number of contacts alleviates more the response when *γ* and *κτ* are small and *h* is large.

## Conclusions

Some conditions must be satisfied for the existence of evenness curtailing of response. First, the expected response must be negligible for small pathogen charges and grow sharply at a certain point. This condition is satisfied *h >* 1 in Fig. 1. The effect is more substantial for large values of *h*, which translates into steeper curves. Second, the ratio between the utmost pathogen charge, *κτ*, and the 50 % response charge, *Q*, must be below a threshold. The lower the fraction of infected people in the population, *γ*, the higher the threshold, represented by the black lines of Fig. 5G-I. Therefore the evenness curtailing is observed in activities where the time spent close to other people is not high, with the population primarily unexposed to the pathogen. People in such situations during an epidemic with *h >* 1, should move around instead of staying too long near the same neighbors.

The present analysis does not encompass the whole dynamics of such a complex phenomenon as the evolution of an epidemic. Nevertheless, it is a tool for understanding specific responses in certain circumstances and clarifying the dynamics’ details. Furthermore, it demonstrates the importance of investigating the precise shape of the dose-response curve and determining the curve concavity, mainly for small pathogen charges.

## Data Availability

The data used in the analysis is in the Supplementary Information.

