## Supplemental text for "When does contacting more people lessen the transmission of infectious diseases?"

### Supporting information

**S1 Appendix Comparing the Hill and the Weibull functions** As discussed in [21] the non-linear dose-response curve is usually described by the Hill function,

$$\pi(q) = \frac{q^h}{Q^h + q^h}, \tag{S1}$$

which we used in this paper, or by the Weibull distribution,

$$\pi_W(q) = 1 - e^{-(q/K)^m}. \tag{S2}$$

These functions can be approximated as $\pi(q) \approx (q/Q)^h$ and $\pi_w(q) \approx (q/K)^m$, respectively for $q \ll Q$ and $q \ll K$. Therefore, by doing $K = Q$ and $m = h$, both functions are indistinguishable for small values of $q$. However, as can be seen in Fig. S1A and S1B, the shapes of the curves are distinct for large values of $q$. Besides, the pathogen charge responsible for 50 % response is $Q$ for the Hill curve and is given by

$$Q_W = K(\ln 2)^{1/m} \tag{S3}$$

for the Weibull distribution.

One way of comparing these curves is by choosing $K$ and $m$ that result in similar behavior around a certain response level $\tilde{\pi}$. This is done by finding the values of $\tilde{q}$, $m$ and $K$ that solve the system of equations

$$\pi(\tilde{q}) = \tilde{\pi} \tag{S4a}$$
$$\pi_w(\tilde{q}) = \tilde{\pi} \tag{S4b}$$
$$\left.\frac{d\pi}{dq}\right|_{\tilde{q}} = \left.\frac{d\pi_w}{dq}\right|_{\tilde{q}}. \tag{S4c}$$

Table S1 contains the solution of these equations for some combinations of $h$ and $\tilde{\pi}$. Figure S1 shows the curves corresponding to the parameters $m$ and $K$ of Table S1.

**Table S1. Parameters of $m$ and $K$ of the Weibull function, Eq. (S2), that touches the Hill function, Eq. (S1), at the point $(\tilde{q}, \tilde{\pi})$, defined by Eq. (S4a-c), for several combinations of $\tilde{\pi}$ and $h$. The value of the half response charge, $Q_W$, is given by Eq. (S3).**

| | $\tilde{\pi} = 0$ | | | $\tilde{\pi} = 0.25$ | | | $\tilde{\pi} = 0.5$ | | | $\tilde{\pi} = 0.75$ | | | $\tilde{\pi} = 0.95$ | | |
|---|---|---|---|---|---|---|---|---|---|---|---|---|---|---|---|
| $h$ | $m$ | $K/Q$ | $Q_W/Q$ | $m$ | $K/Q$ | $Q_W/Q$ | $m$ | $K/Q$ | $Q_W/Q$ | $m$ | $K/Q$ | $Q_W/Q$ | $m$ | $K/Q$ | $Q_W/Q$ |
| 0.25 | 0.25 | 1.00 | 0.23 | 0.22 | 3.82 | 0.71 | 0.18 | 7.63 | 1.00 | 0.14 | 7.24 | 0.48 | 0.08 | 0.13 | 0.00 |
| 0.50 | 0.50 | 1.00 | 0.48 | 0.43 | 1.95 | 0.84 | 0.36 | 2.76 | 1.00 | 0.27 | 2.69 | 0.69 | 0.16 | 0.36 | 0.04 |
| 0.70 | 0.70 | 1.00 | 0.59 | 0.61 | 1.61 | 0.88 | 0.50 | 2.07 | 1.00 | 0.38 | 2.03 | 0.77 | 0.22 | 0.48 | 0.09 |
| 1.00 | 1.00 | 1.00 | 0.69 | 0.87 | 1.40 | 0.92 | 0.72 | 1.66 | 1.00 | 0.54 | 1.64 | 0.83 | 0.32 | 0.60 | 0.19 |
| 1.40 | 1.40 | 1.00 | 0.77 | 1.22 | 1.27 | 0.94 | 1.01 | 1.44 | 1.00 | 0.76 | 1.42 | 0.88 | 0.44 | 0.69 | 0.30 |
| 2.00 | 2.00 | 1.00 | 0.83 | 1.74 | 1.18 | 0.96 | 1.44 | 1.29 | 1.00 | 1.08 | 1.28 | 0.91 | 0.63 | 0.77 | 0.43 |
| 4.00 | 4.00 | 1.00 | 0.91 | 3.48 | 1.09 | 0.98 | 2.89 | 1.14 | 1.00 | 2.16 | 1.13 | 0.96 | 1.27 | 0.88 | 0.66 |

Although it is possible to tune the Weibull function to make it behave like the Hill function over a vicinity of $\tilde{q}$, it is impossible to superpose the functions over the whole range of $q$ due to their distinct shape. For this reason, both functions may fit a narrow data set, but both functions cannot simultaneously describe a broad enough data set.

Even if the curves are indistinguishable around the narrow data set they fit, they will have a distinct behavior in other regions. For example, if the data are clustered in

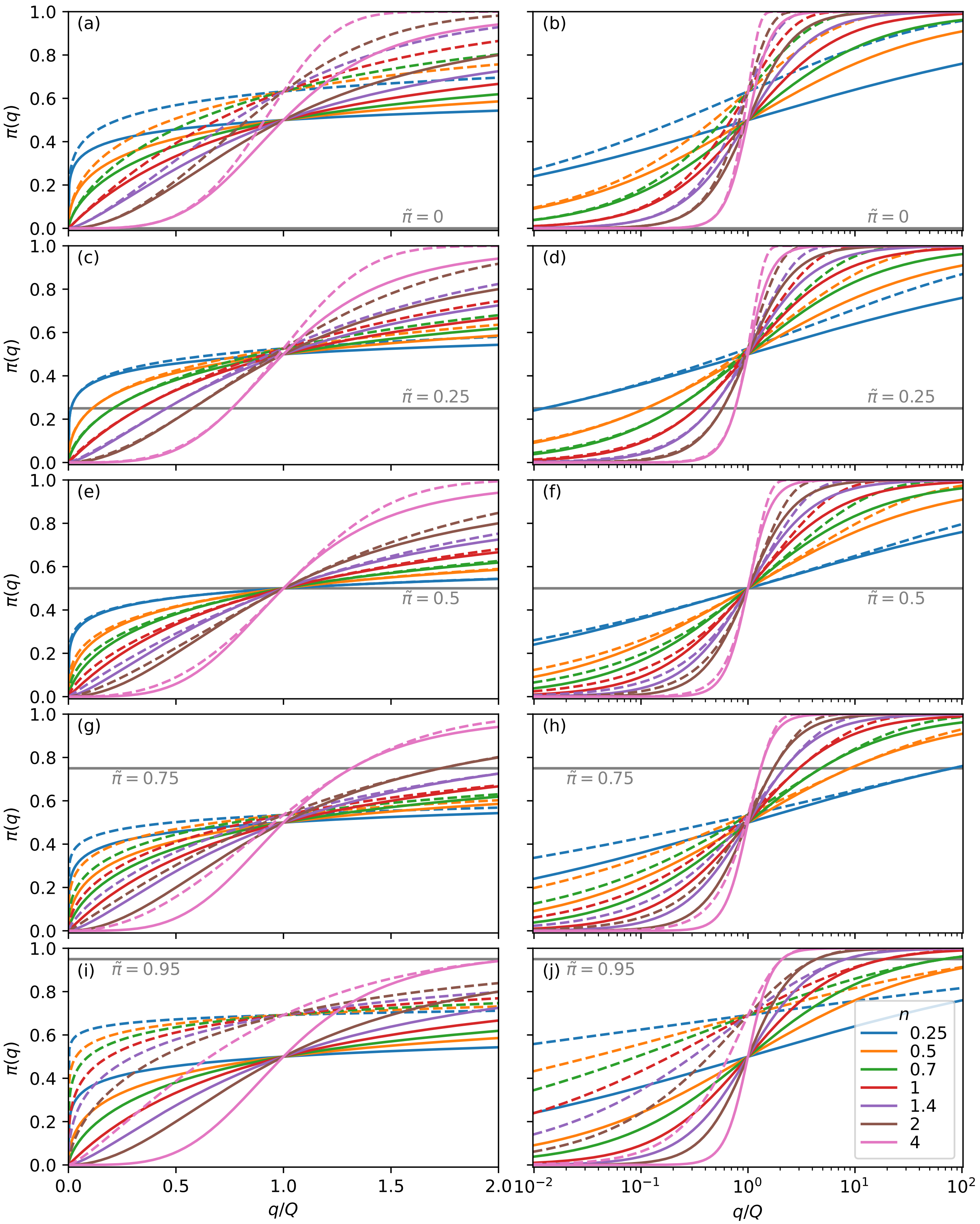


**Fig S1. Comparing the Hill and Weibull curves.** The Hill (Eq. (S1), continuous lines) and the Weibull (Eq. (S2), dashed lines) curves corresponding to the parameters $h$, $m$, and $K$ from Table S1. The $x$-axes of the plots at the left and right sides are, respectively, linear and logarithmic.

the vicinity of $\tilde{\pi} \sim 0.95$, Fig. S1(i),(j), the Hill curve and the Weibull will behave distinctly for small values of $q$. Therefore, data with large values of $\pi$ provide no information about the behavior of the curve for small values of $\pi$ if the exact form of the curve is not known, and vice-versa.

On the other hand, if the data is clustered around a small value of $\tilde{\pi}$, the Hill and Weibull curves behave similarly, as can be deduced from Fig. S1A-C and the identical approximations valid for $q \to 0$. In these conditions, both curves provide the same results when describing phenomena involving small values of $q$, as is the case of low exposition. For example, Table S1 shows that when $\tilde{\pi} \sim 0.25$, the difference between $m$ and $h$ and between $Q$ and $Q_W$ are slight, and the curves are very similar in the interval $0 \leq q \leq Q$ in Fig. S1C-D.

**S2 Appendix The experimental values of the curve parameters** In this section, we review previous works on response curves for several pathogens and outcomes. The goal is to provide an outlook instead of exhaustive coverage of the subject. For each of the studies presented in this section, we fitted Eq. (5) and plotted the corresponding curve along with the experimental data. The legends display the values of the curves' parameters $h$ and $Q$. When adjusting the parameters, special attention was paid to the low doses part of the curve ($\pi \lesssim 0.5$), since the behavior of the curve at this region is responsible for the reduced response when $N_c$ is increased with $h > 1$.

Measuring pathogen doses is often tricky, particularly for human diseases, when controlled contamination is usually unacceptable. For this reason, most works in this direction involve mathematically modeling the dispersion of pathogen as a function of the subjects' behavior. Thus, the works discussed here involve some mathematical modeling with the exception of [3].

MCKenney, Kurath and Wargo investigated two strains of the infectious hematopoietic necrosis virus (IHNV) with distinct fitness and virulence [3]. The ability to infect a host was determined by exposing juvenile rainbow trout hosts to a wide range of virus doses. The infectivity data obtained by them is shown in Fig. S2.

A model that captures the dose-timing pattern was introduced by Mayer *et al.* in [4] to investigate the empirical time-series data of inhalational anthrax in monkeys. The model captures the time dependence in a manner that incorporates the immune response dynamics to the inhalation data presented by Brachman *et al.* in [5]. In Fig. S3 we reproduce the plots that summarize their results.

In [8], Schiffer *et al.* developed a mathematical model based on reproducing shedding patterns in transmitting partners to estimate the infectivity of single viral particles. The model was used to inferred probability estimates for transmission at different levels of genital tract viral load in the transmitting partner resulting in the data reproduced in Fig. S4.

To assess the risk of transmitting human herpervirus 6 to infants, the virus shedding by mothers and other children cohabiting the house was modeled by Mayer *et al.* in [7]. These estimates were used to produce the plots shown in Fig. S5.

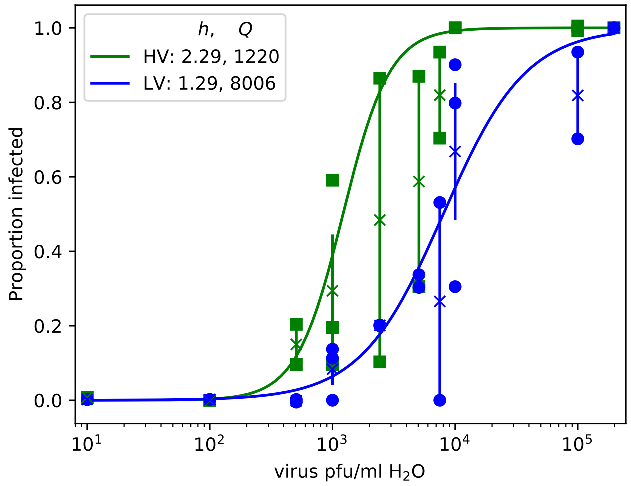


**Fig S2. Parameter $h$ from [3].** Data from [3] relating the probability of infection to the doses of IHNV received. The green squares and the blue circles refer to the “high virulence” and “low virulence” strains. From these points, we calculated the mean value and the error plotted as crosses with error bars. The lines are fittings with Eq. (5), with the parameters shown in the legend.

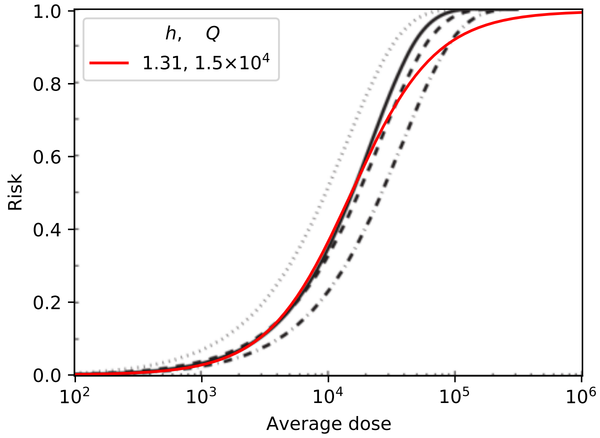


**Fig S3. Parameter $h$ from [4].** Reproduction of Fig. 5 of [4], which presents the best-fit of several models to inhalation anthrax mortality data in monkeys [5]. The red line is Eq. (5) with parameters in the legend, which presents a good agreement with the curves at the small values of $\pi$.

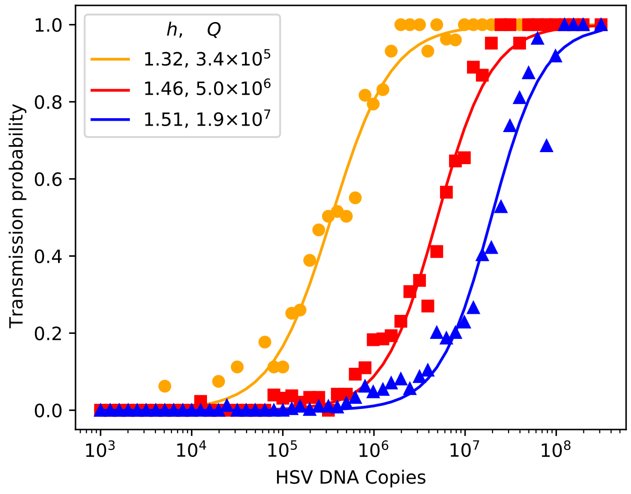


**Fig S4. Parameter $h$ from [8].** Data points from [8], describing the transmission probability of herpes simplex virus-2 as a function of the quantity of viral shedding. Simulations were performed with different values for the parameter describing the infectivity: high infectivity (orange circles), medium infectivity (red squares), and low infectivity (blue triangles). The lines are the fitting of Eq. (5) with the parameters shown as legends.

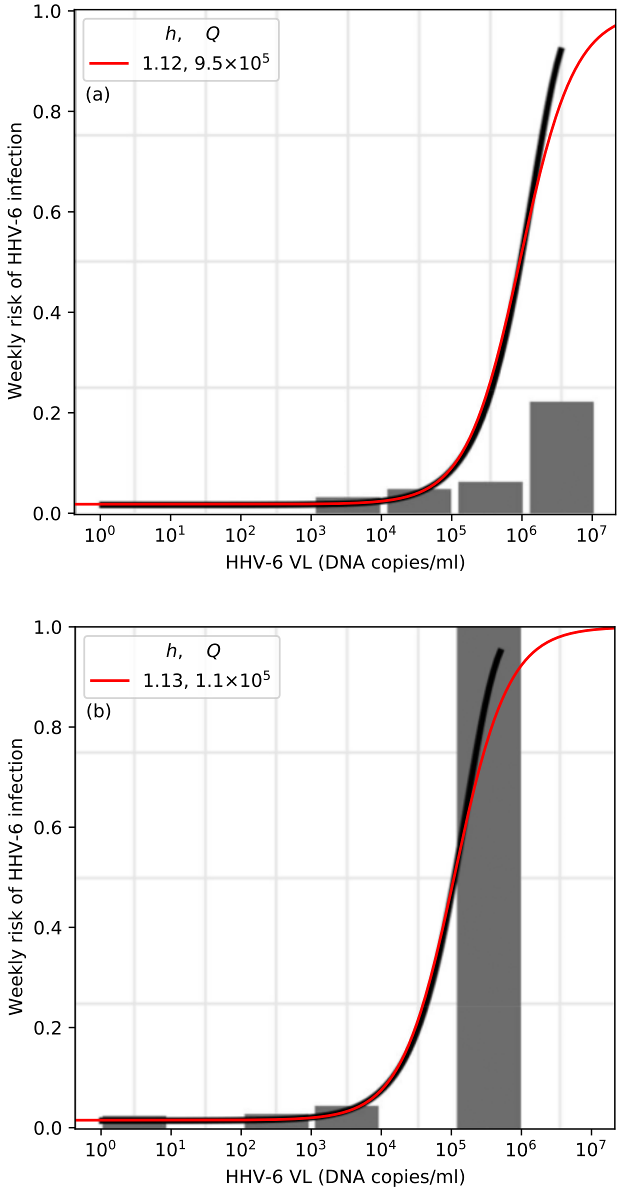


**Fig S5. Parameter $h$ from [7].** Reproduction of Fig. 3 of [7], which presents the estimation of human herpesvirus 6 acquisition risk from exposures by week using a combined exposure model. The infant was exposed to the virus shedding by an infected mother or a secondary child living in the same home. In both plots, the black lines depict estimated risk from the model, and the red lines are Eq. (5) with the parameters adjusted to present a good agreement for small values of $\pi$. Risk of an infant being infected by (a) a secondary child or (b) the mother.

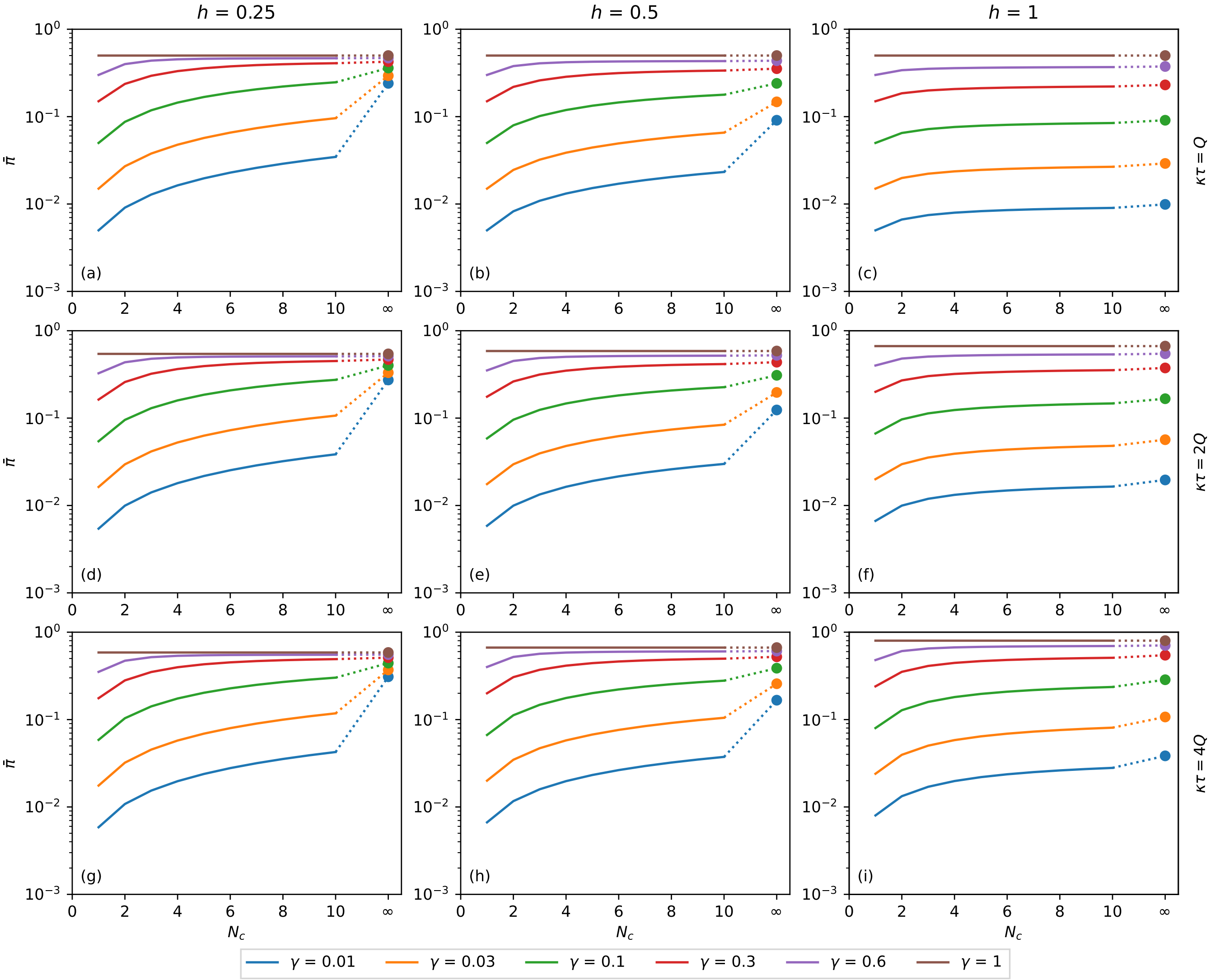


**Fig S6. Binomial distribution with $h \leq 1$.** Binomial expected response as a function of $N_c$. It is calculated as a function of the number of contacts for the binomial distribution of pathogen charge, Eq. (10)-(11) applied to Eq. (8). Although $N_c$ is an integer variable, the functions are shown as lines to make the plots less bulky. The dashed vertical lines are the points of maximum predicted by the approximation Eq. (14). The hollow circles are the points of maximum of each combination of $h$, $\kappa\tau$, and $\gamma$. The filled circles are the values of $\bar{\pi}$ at $N_c \to \infty$, calculated as $\pi(\gamma\kappa\tau)$, Eq. (15). (a) $\kappa\tau = Q$, $h = 0.25$, (b) $\kappa\tau = Q$, $h = 0.5$, (c) $\kappa\tau = Q$, $h = 1$, (d) $\kappa\tau = 2Q$, $h = 0.25$, (e) $\kappa\tau = 2Q$, $h = 0.5$, (f) $\kappa\tau = 2Q$, $h = 1$, (g) $\kappa\tau = 4Q$, $h = 0.25$, (h) $\kappa\tau = 4Q$, $h = 0.5$, (i) $\kappa\tau = 4Q$, $h = 1$.
